# LLM-Driven Target Trial Emulation with Human-in-the-Loop Validation for Randomized Trial: Automated Protocol Extraction and Real-World Outcome Evaluation^Ψ^

**DOI:** 10.64898/2026.04.09.26350523

**Authors:** Samrat Kumar Dey, Adnan I. Qureshi, Chi-Ren Shyu

**Affiliations:** Institute for Data Science and Informatics, University of Missouri, Columbia, MO, USA; Department of Neurology, University of Missouri, Columbia, MO, USA; Zeenat Qureshi Stroke Institute, USA; Department of Electrical Engineering and Computer Science, University of Missouri, Columbia, MO, USA

## Abstract

Target trial emulation (TTE) enables causal inference from observational data but remains bottlenecked by manual, expert-dependent protocol operationalization. While large language models (LLMs) have advanced clinical knowledge extraction and code generation, their ability to automate end-to-end TTE workflows remains largely unexplored. We present an LLM-driven framework using retrieval-augmented generation to extract the five core TTE design parameters from the Carotid Revascularization and Medical Management for Asymptomatic Carotid Stenosis Trial (CREST-2) protocol and generate executable phenotyping pipelines for real-world EHR data. The performance of the framework was evaluated along two dimensions. First, protocol extraction accuracy was assessed against a gold-standard checklist of trial design components using precision, recall, and F1-score metrics. Second, outcome validity was evaluated through population-level concordance analyses comparing EHR-derived outcomes with published trial endpoints using standardized mean difference, observed-to-expected ratios, confidence interval overlap, and two-proportion z-tests. Further, Human-in-the-loop validation assessed the correctness of extracted clinical logic and phenotype definitions. Together, these evaluations demonstrate a structured approach for assessing LLM-driven protocol-to-pipeline translation for scalable real-world evidence generation.

## Introduction

Producing reliable causal evidence from real-world healthcare data remains a major challenge in clinical and translational research. Although randomized controlled trials (RCTs) are the gold standard for assessing treatment effects, they are often difficult to conduct because of ethical concerns, high costs, limited generalizability, and restrictive patient eligibility^1^. As a result, researchers are increasingly turning to observational data sources, especially electronic health records (EHRs) to generate real-world evidence (RWE). This evidence helps support clinical decision-making and health-policy decisions^2,3^. Although EHR-based data provide large amounts of detailed clinical information, leveraging them for causal inference remains methodologically challenging^4–6^.

Target trial emulation (TTE) has emerged as a powerful approach for addressing many of these limitations^7,8^. Pioneered by Hernán *et al*.^9^, TTE requires investigators to define the protocol of a target trial in detail. This includes eligibility criteria, treatment strategies, treatment assignments, follow-up periods, outcomes, causal contrasts, and statistical analysis plans, which are then systematically emulated using available observational data^10^. Its use has grown rapidly across diverse therapeutic areas, including cardiovascular disease, oncology, nephrology, and infectious disease^11–13^. The recent publication of the TARGET (Transparent Reporting of Observational Studies Emulating a Target Trial)^14^ by Cashin *et al*. reporting guideline highlights the growing maturity and broad acceptance of target TTE as a standard for rigorous observational research. Despite its methodological rigor, TTE remains an expert-driven, resource-intensive process in practice^15^. Designing a valid emulation requires substantial multidisciplinary expertise across clinical medicine, epidemiology, biostatistics, and causal inference theory^16^. The current human reliant pipeline for causal analysis, while essential, remains slow, error prone and limited by the availability of specialized expertise, which restricts both the throughput and accessibility of high quality TTE studies^17^.

Recent advances^18^ have shown that large language models (LLMs) possess promising capabilities in causal reasoning, achieving high accuracy in pairwise causal discovery, counterfactual reasoning, and event causality tasks, often outperforming traditional computational methods^19^. In clinical settings, LLMs have shown promise for automated patients to trial matching, eligibility criteria extraction, clinical data processing, and study protocol generation ^20–22^. Critically, emerging work has proposed conceptual frameworks LLMs as “causal copilots”-AI driven assistants that encode cross domain knowledge and can engage researchers through natural language interactions to support study design, identify potential biases, and guide the specification of causal estimands^23,24^. Despite the well-established methodological foundation of target trial emulation and the rapidly advancing capabilities of LLMs in healthcare, a critical gap remains at their intersection. Current TTE implementations require substantial human expertise at every stage, from formulating the causal question and specifying the target trial protocol to identifying confounders, mapping variables from EHR data, and selecting appropriate statistical methods. This expertise heavy workflow creates significant bottlenecks: many causal questions that could be answered using available observational data remain unaddressed, simply because the required multidisciplinary teams are unavailable or the manual design process is comparatively slow^23^. Meanwhile, LLMs are being applied to isolated aspects of clinical research, such as literature synthesis, cohort identification, and criteria extraction. However, these applications lack a unified framework that integrates them into the end-to-end causal inference pipeline that target trial emulation requires^20,21,24^. The absence of an integrated, LLM-driven approach to target trial emulation represents a missed opportunity.

This study hypothesizes that LLMs, when combined with retrieval from clinical trial protocols and integrated into a structured human-supervised workflow, can automatically extract and operationalize the key design components of a TTE. These components include eligibility criteria, treatment definitions, time zero, outcomes, and follow-up rules. Using these extracted elements, the automated pipeline can generate real-world outcome estimates from EHR data that are clinically plausible and consistent with published trial results. We further hypothesize that this approach shifts the role of the clinical informatician from manually constructing analytic pipelines to validating automatically generated workflows, providing a scalable foundation for future causal inference studies. This research work is guided by the following research questions **(RQ)**:

### RQ1

To what extent can LLMs automatically extract and operationalize valid target trial emulation design parameters such as eligibility criteria, treatment strategies, time-zero assignment, outcomes, causal contrasts, and follow-up periods from a published clinical trial protocol and structured data dictionaries, in accordance with established TTE methodological standards?

### RQ2

How do the real-world outcome estimates produced by the LLM-driven target trial emulation, when applied to EHR data, compare with the published efficacy and safety endpoints from the CREST-2 randomized clinical trial?

This work makes the following contributions **(C)** to the fields of clinical informatics and real-world evidence generation:

### C1

We developed an LLM-driven framework for automated target trial emulation, where a LLM parses a clinical trial protocol and extracted the five key design parameters such as eligibility, treatment strategy, time zero, outcomes, and follow-up to generate an executable analytic pipeline. (RQ1)

### C2

We provided a systematic evaluation of human-in-the-loop validation, identified where LLM-generated clinical logic requires expert review and demonstrated that domain oversight remained essential for ensuring clinically valid phenotyping and code definitions. (RQ1)

### C3

We applied the framework to real-world EHR data and compared the resulting outcome estimates with published randomized trial endpoints, demonstrating that an LLM-driven emulation pipeline could generate clinically plausible and interpretable real-world evidence. (RQ2)

## Methods

### Study Design and Overview

We developed and compared an LLM-driven pipeline for TTE of the Carotid Revascularization and Medical Management for Asymptomatic Carotid Stenosis Trial (CREST-2)^25^. The study comprised two phases corresponding to our two research questions. In the first phase (RQ1), a LLM was tasked with autonomously extracting and operationalizing the five core TTE design parameters (eligibility, treatment, time-zero, outcome, and follow-up) from the published CREST-2 protocol. In the second phase (RQ2), the resulting pipeline was applied to real-world electronic health record (EHR) data to generate outcome estimates, which were then compared against the published CREST-2 trial endpoints. The CREST-2 trial^25^, published by Brott *et al*. in the New England Journal of Medicine in 2025, comprised two parallel, observer-blinded randomized clinical trials conducted at 155 centers across five countries. One trial compared carotid artery stenting (CAS) plus intensive medical management (IMM) versus IMM alone; the other compared carotid endarterectomy (CEA) plus IMM versus IMM alone. Each trial enrolled over 1,200 patients with high-grade (≥70%) asymptomatic carotid stenosis. The primary outcome was a composite of any stroke or death within 44 days of randomization, or ipsilateral ischemic stroke during the remaining follow-up period up to four years.

### Proposed Target Trial Emulation (TTE) Framework

We adopted the target trial emulation framework as described by Hernán *et al*.^9^, which specifies that an observational study should explicitly emulate a hypothetical randomized trial by defining five core design parameters (DPs): (1) eligibility criteria, (2) treatment strategies, (3) time-zero assignment, (4) outcome definitions, and (5) follow-up and censoring specifications. Each parameter was extracted from the CREST-2 protocol by LLM and mapped to executable analytic logic against the ORACLE EHR Healthcare Data Model^26^. **Figure 1** illustrates the end-to-end system architecture of the proposed LLM-driven target trial emulation framework. The architecture comprises three core LLM-driven modules, two human-in-the-loop validation roles, and two downstream execution pipelines.

**Figure 1.**
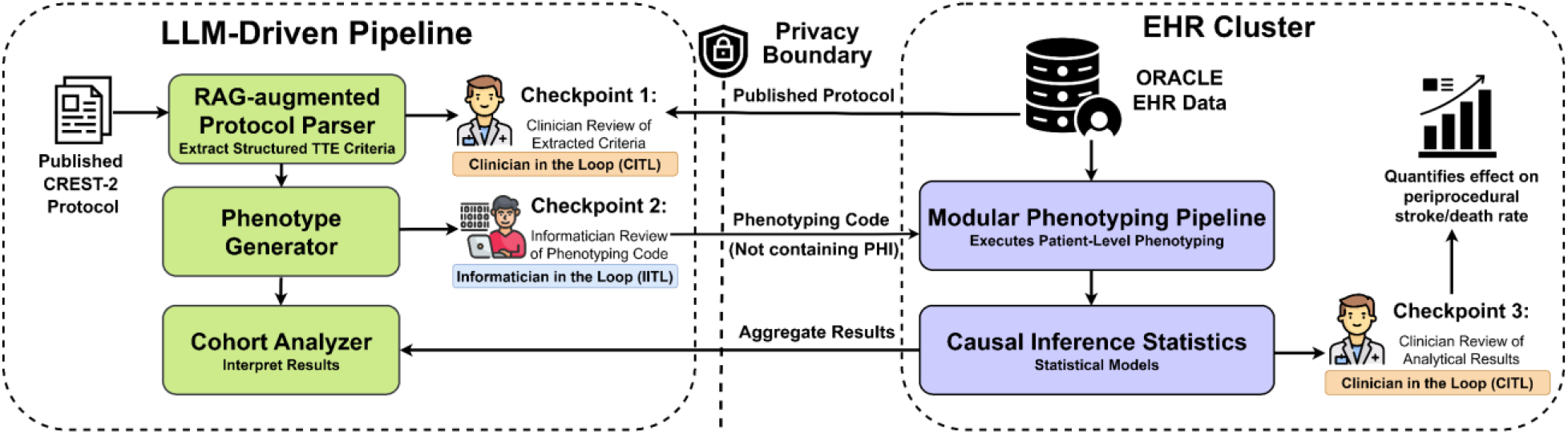
System Architecture for LLM-Driven Target Trial Emulation.

The framework operates through three sequential modules. The Retrieval-Augmented Generation (RAG)-augmented protocol parser ingests the published clinical trial protocol^25^ and structured data dictionaries, using retrieval-augmented generation to extract the five-target trial emulation design parameters into a structured intermediate representation. The Phenotype Generator translates these extracted parameters into executable computable phenotype definitions, mapping clinical concepts to standardized terminology such as International Classification of Diseases, 10^th^ Revision, Clinical Modification (ICD-10-CM), Current Procedural Terminology (CPT) and generating code logic against the EHR data model. The cohort analyzer interprets the resulting cohort characteristics and outcome distributions, providing summary statistics, validation checks, and diagnostic reporting.

Two human-in-the-loop roles were included to ensure both clinical and technical accuracy. The Clinician in the Loop (CITL) reviews the extracted protocol parameters to confirm that eligibility criteria, outcome definitions, and clinical code set correctly represent the original trial protocol. The clinician also evaluates the final statistical results to ensure they are clinically reasonable. The Informatician in the Loop (IITL) reviews the technical implementation of the pipeline, including phenotype logic, data model mappings, and generated code, to ensure that the computational workflow correctly follows the clinical specifications. The validated outputs feed into two downstream execution modules. The Modular Phenotyping Pipeline applies the LLM-generated phenotype definitions to patient-level EHR data, executing cohort assembly, outcome classification, and event ascertainment at scale. The Causal Inference Statistics module performs the formal statistical analyses to produce the outcome estimates for comparison against published trial endpoints.

### Data Source

This study used Oracle Health Real-World Data (OHRWD)^26^, a deidentified, EHR-derived database from US health care facilities that maintain data use agreements with Oracle Corporation, to perform LLM-driven target trial emulation. We analyzed the expert determination release “real_world_data_ed_feb_2025.” The February 2025 OHRWD release includes data from 149 US health systems, with representation across US Census regions (South 29%, West 27%, Midwest 24%, Northeast 20%)^26^. The study period ran from January 1, 2015, through February 28, 2025, and covered longitudinal inpatient and outpatient encounters, procedures, diagnoses, and mortality data. As part of deidentification, facility identifiers and patient geographic location were not available. As the dataset is deidentified, institutional review board review and informed consent were not required.

### LLM-Driven Protocol Parsing and Code Generation Model and Prompting Strategy

**Figure 2** presents the five-module sequential pipeline that operationalizes the LLM-driven target trial emulation. Each module has explicitly defined inputs and outputs, creating an auditable chain from the raw trial protocol to outcome estimates.

**Figure 2.**
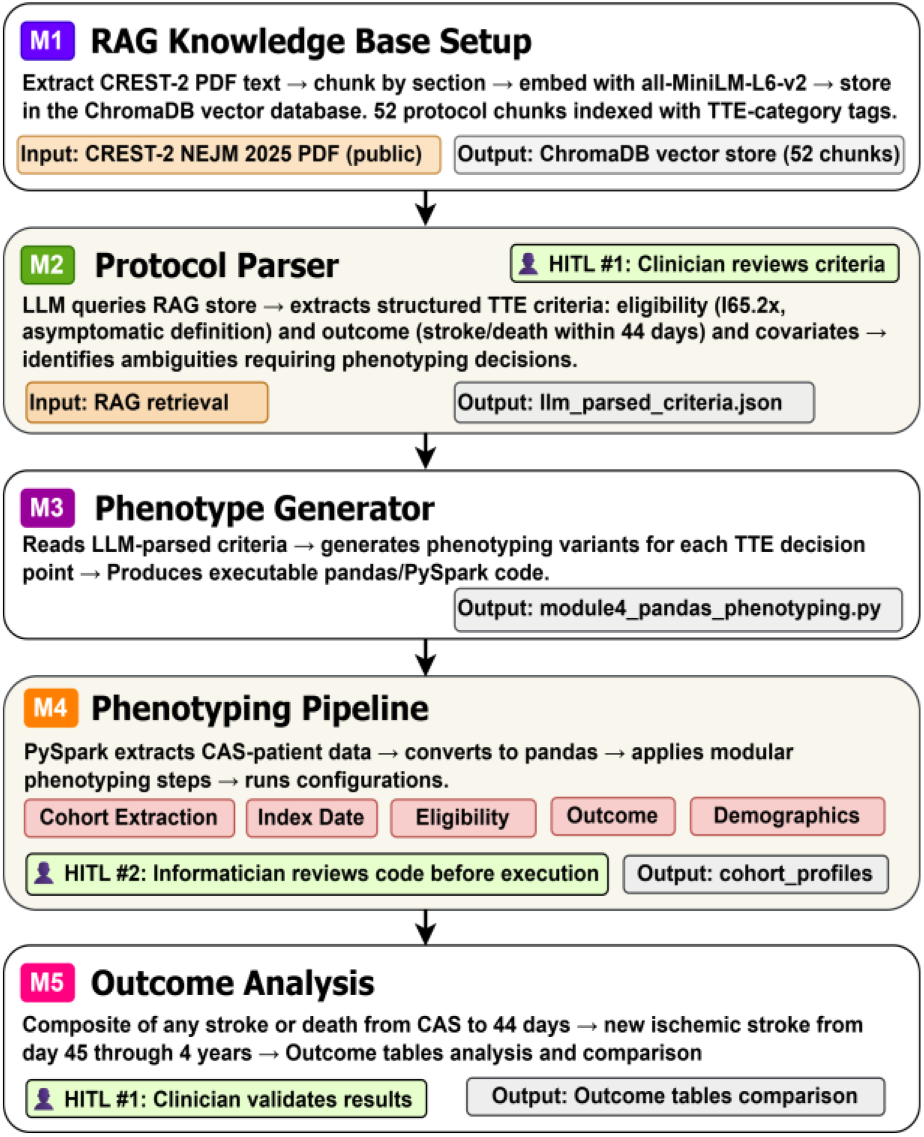
Modular Pipeline Workflow for LLM-Driven Target Trial Emulation.

For pipeline execution and phenotyping tasks that involve direct interaction with patient-level EHR data, we deployed Qwen3:8B^27^ locally via Ollama^28^ to ensure that no protected health information (PHI) was transmitted to external servers. Ollama provides a local inference runtime that executes the language model entirely on-premises, eliminating network-based data exposure. The Qwen3:8B model^27^ was selected based on its demonstrated capabilities in medical reasoning and clinical NLP tasks. This approach ensures compliance with HIPAA and institutional data governance policies while maintaining the analytical quality needed for clinical research.

Module 1 (M1: RAG Knowledge Base Setup) establishes the retrieval-augmented generation infrastructure. The published CREST-2 protocol is extracted, segmented into 52 text chunks organized by TTE-category tags, embedded using the all-MiniLM-L6-v2^29^ sentence transformer model, and stored in a ChromaDB^30^ vector database.

Module 2 (M2: Protocol Parser) performs the core knowledge extraction task. The LLM queries the RAG store to retrieve relevant protocol sections and extracts structured TTE criteria, including eligibility definitions (e.g., I65.2x for carotid stenosis, asymptomatic definition via absence of ipsilateral stroke/TIA within 180 days), outcome specifications (composite of stroke/death within 44 days, ipsilateral ischemic stroke thereafter), and covariate definitions. At this stage, Human-in-the-Loop checkpoint #1 (HITL #1) engages a clinician to review the extracted criteria for clinical accuracy and completeness before they are passed to the code generation module.

Module 3 (M3: Phenotype Generator) translates the parsed criteria into executable analytic code. Using the applied LLM, the module reads the structured JSON output from M2 and generates phenotyping variants for each TTE decision point including ICD-10-CM code sets for anterior and posterior circulation stroke, CPT procedure codes for CEA and CAS, temporal logic for follow-up windows and blanking periods, and encounter-type filters.

Module 4 (M4: Phenotyping Pipeline) executes the generated code against patient-level EHR data within the institutional computing environment. Any LLM-assisted tasks at this stage such as code debugging, query optimization, or result interpretation use Qwen3:8B running locally via Ollama, ensuring that no patient data is transmitted to external servers. Human-in-the-Loop checkpoint #2 (HITL #2) engages an informatician to review the generated code for technical correctness verifying data model mappings, join logic, and temporal filters before the pipeline is executed against real world data.

Module 5 (M5: Outcome Analysis) performs the statistical analyses that produce the final study outputs, with all computations executed locally. The module computes the composite endpoint of any stroke or death from the index procedure through 44 days (periprocedural window) and new ischemic stroke from day 45 through 4 years (long-term window). Human-in-the-Loop checkpoint #1 (HITL #1) re-engages the clinician to validate the clinical plausibility of the results assessing whether event rates, and outcome distributions are consistent with clinical expectations for a carotid revascularization population.

A structured prompting strategy was employed, with separate prompts for each of the five design parameters. Each prompt specified the clinical context, the target parameter to extract, the output format and any constraints including inpatient encounters only, specific ICD-10/CPT code sets and others.

### Design Parameter (DP-1): Eligibility Criteria

The LLM was prompted to define inclusion and exclusion criteria for CREST-2 emulation. Inclusion criteria required: age ≥18 years at index procedure date; (b) high-grade asymptomatic carotid stenosis (≥70%); (c) receipt of a qualifying CEA or CAS procedure identified by CPT (CPT) codes; and (d) at least 1 year of continuous enrollment in the data source prior to the index date. Exclusion criteria included: (a) ipsilateral stroke or transient ischemic attack (TIA) within 180 days before the index procedure ensuring an asymptomatic population; (b) prior ipsilateral CEA or CAS before the index date; and (d) missing critical demographic data (sex, birth date). The LLM generated phenotyping code implementing these criteria against the ORACLE EHR DEMOGRAPHIC, PROCEDURES, DIAGNOSIS, and ENCOUNTER tables. **Figure 3** (A) shows the natural-language prompt with the inclusion/exclusion criteria on the left and the LLM-generated coding phenotype with CPT/ICD filters on the right. It shows that the LLM correctly translated the (1) code sets and (2) 180-day lookback.

**Figure 3.**
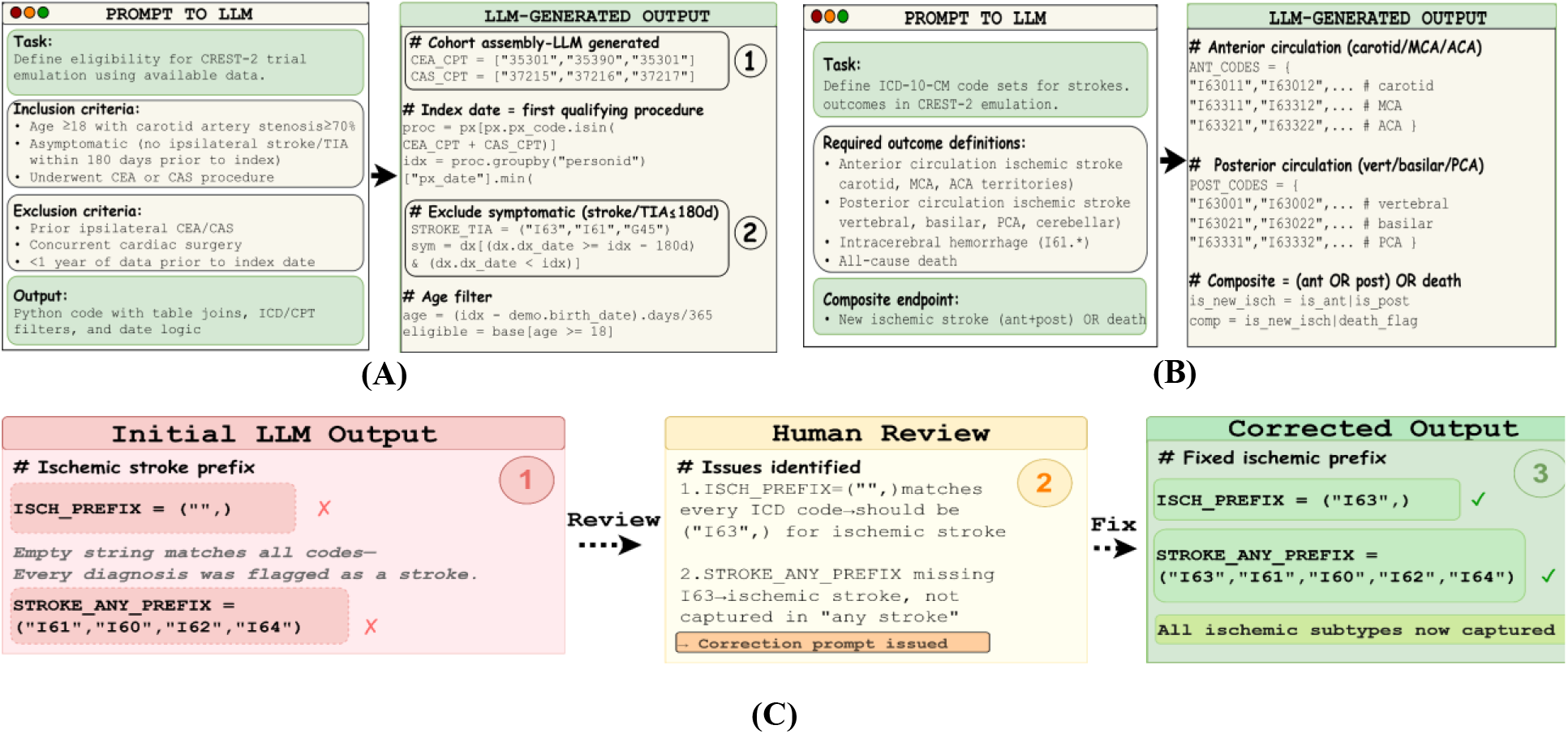
Representative prompts and corresponding LLM-generated outputs across three pipeline stages.

### Design Parameter (DP-2): Treatment Strategies

Treatment assignment was defined by the index procedure, patients undergoing carotid endarterectomy (CEA) comprised one arm, and patients undergoing carotid artery stenting (CAS) comprised the other. The index date was defined as the date of the first qualifying revascularization procedure. In this analysis, we reported results across CAS treatment arms, as the primary objective was to evaluate the LLM pipeline’s ability to generate valid outcome estimates rather than to estimate treatment effects.

### Design Parameter (DP-3): Time-Zero Assignment

Time zero (T^0^) was defined as the date of the first qualifying CAS procedure for each patient. This aligns with the CREST-2 trial design, where randomization occurred at the time of treatment assignment. The follow-up window was divided into two windows consistent with the CREST-2 primary endpoint structure: a periprocedural window from day 0 through day 44, and a long-term efficacy window from day 45 through 4 years (1,460 days) post-index.

### Design Parameter (DP-4): Outcome Definitions

**Figure 3** (B) shows the prompting and generated ICD-10-CM code sets for stroke outcomes classified by vascular territory, based on the anatomical descriptions in the CREST-2 protocol. The ascertained of outcomes were similar to the CREST-2 endpoints and were evaluated in two distinct, non-overlapping windows, (1) stroke or death within 44 days mirroring the peri-procedural outcome window used in CREST-2; and (2) new ischemic stroke from day 44 up to 4 years as post-procedural follow-up. This study also investigates (i) anterior circulation ischemic stroke, (ii) posterior circulation ischemic stroke, (iii) any deaths, (iv) intracerebral hemorrhage (ICH) for the observed two endpoints.

### Design Parameter (DP-5): Follow-up and Censoring

The follow-up process was divided into two windows: (1) periprocedural, from T^0^ to T^0^+ 44 days; and (2) long-term, from T^0^ + 45 days to T^0^ + 4 years (1,460 days). Patients were censored at the earliest of (a) the end of the observation window (day 44 or day 1,460); (b) the last known encounter date (loss to follow-up); or (c) the end of data availability. Time-to-event was computed as the number of days from the index date to the first composite event, or to the censoring date for event-free patients.

### Human-in-the-Loop Validation

All LLM-generated code and design parameter specifications underwent structured reviews by a clinical domain expert. Human-in-the-loop (HITL) validation in this pipeline is necessary because target trial emulation requires precise clinical logic and coding definitions, where small errors in code sets or temporal rules can lead to clinically incorrect cohort construction and outcome identification. This HITL validation functions as a quality assurance layer that ensures clinical validity while allowing the LLM to automate the majority of the protocol-to-pipeline translation process. The review process followed a systematic protocol: (1) verification of ICD-10-CM and CPT code set accuracy against published coding references; (2) logical validation of inclusion/exclusion criteria implementation; (3) verification of temporal logic (date comparisons, window boundaries, blanking periods); and (4) execution testing with diagnostic output to confirm expected counts and distributions. During this review as shown in **Figure 3** (C), specific errors were identified and corrected. In the figure it shows, the LLM initially defined the ischemic stroke prefix as an empty string (ISCH_PREFIX = (““,)), which would match all ICD-10 codes rather than only I63.* codes. Additionally, the initial stroke-any definition omitted I63 from the prefix list, excluding all ischemic strokes from the broad stroke category. These errors were corrected through an iterative prompting and review cycle.

### Prompting Techniques Mapped to Pipeline Modules

We implemented a modular pipeline in which different prompting strategies were applied as shown in **Figure 4** across workflow modules. The protocol parsing and phenotype generation modules (M2–M3) used retrieval-augmented generation (RAG), schema-constrained structured output prompting, and task decomposition to extract target trial emulation design parameters and generate executable code from the CREST-2 protocol. The downstream execution and analysis modules (M4–M5) employed chain-of-thought reasoning, multi-turn iterative refinement, and domain-constrained prompting to validate phenotype logic, execute the EHR analysis pipeline, and produce outcome estimates.

**Figure 4.**
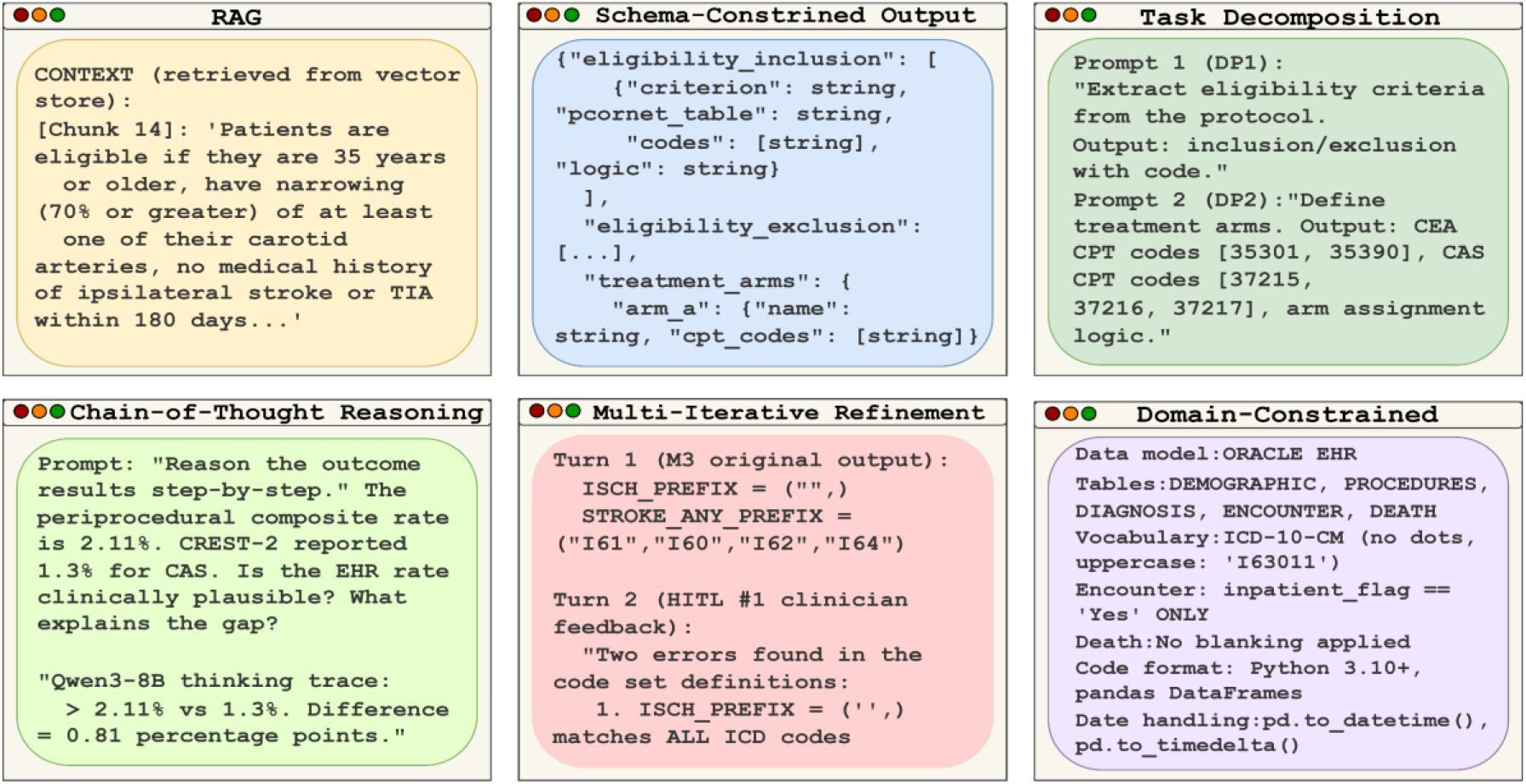
Prompting strategies used in the LLM driven pipeline for target trial emulation.

## Results

### Baseline Characteristics

The analytic cohort included 5,831 patients presented in **Table 1** who underwent CAS. To enable direct comparison with the randomized trial, we report only baseline demographic and clinical characteristics that were also presented in the CREST-2 trial publication. The mean (SD) age in the EHR cohort was 72.18 (8.48) years, compared with 69.03 (8.1) years in the CREST-2 cohort. Women comprised 2,162 patients (37.07%) in the EHR cohort, closely matching the 36.9% reported in CREST-2. The majority of patients were White (86.84%), followed by Black or African

**Table 1.** Characteristics of the asymptomatic patients who underwent CAS.

| Variables | EHR (N=5831) | CREST-2 (N=616) |
| --- | --- | --- |
| Age-yr | 72.18 ± 8.48 | 69.03 ± 8.1 |
| **Sex** |  |  |
| Female | 2162 (37.07%) | 227 (36.9%) |
| **Race** |  |  |
| White | 5064 (86.84%) | 572 (92.9%) |
| Black or African | 228 (3.91%) | 35 (5.7%) |
| Other | 477 (8.18%) | 9 (1.5%) |
| **Risk Factors** |  |  |
| Smoking | 860 (14.75%) | 116 (18.8%) |
| Hypertension | 3588 (61.53%) | 542 (88.0%) |
| Dyslipidemia | 2657 (45.57%) | 567 (92.0%) |
| Diabetes mellitus | 1710 (29.33%) | 251 (40.7%) |
| Cerebrovascular disease | 390 (6.69%) | 331 (53.7%) |

American (3.91%) and other races (8.18%), whereas the CREST-2 cohort reported a higher proportion of white participants (92.9%). Regarding cardiovascular risk factors, the EHR cohort demonstrated lower prevalence of hypertension, dyslipidemia, diabetes mellitus, smoking, and cerebrovascular disease compared with the trial population.

### Outcomes at 0–44 Days

**Table 2** presents the LLM-derived outcome estimates for both follow-up windows. During the periprocedural period (0-44 days post-CAS), the composite rate of new ischemic stroke or death was 2.11% (95% CI, 1.76–2.51; 123 events among 5,831 patients). New ischemic stroke of any type occurred in 74 patients (1.27%; 95% CI, 1.00– 1.59), with anterior circulation strokes predominating (36 events; 0.62%) and posterior circulation strokes rare (1 event; 0.02%). Intracerebral hemorrhage occurred in 11 patients (0.19%; 95% CI, 0.09–0.34), and 56 deaths (0.96%; 95% CI, 0.73–1.25) were recorded. In the CREST-2 trial, the periprocedural stroke and death rate in the CAS group was 1.3% with 8 events (7 strokes and 1 death among 616 patients). The LLM-derived EHR estimate of 2.11% is higher than the trial outcome, consistent with expectations for a real-world population that includes operators across a broader range of experience levels and patients who may not meet the stringent anatomical and operator credentialing requirements of CREST-2. Additionally, the EHR composite captures all strokes, not only ipsilateral, which may contribute to the higher rate.

**Table 2.** Outcomes from day 0 to 44 and 45 through 4 years after carotid artery placement on EHR data.

| Outcomes | 0–44 Days Post-CAS Outcome | 45d–4years Post-CAS Outcome |
| --- | --- | --- |
| New ischemic stroke (any) | 74 (1.27%; 95% CI, 1.00–1.59) | 179 (3.07%; 95% CI, 2.64–3.55) |
| Anterior circulation ischemic stroke | 36 (0.62%; 95% CI, 0.43–0.85) | 34 (0.58%; 95% CI, 0.40–0.81) |
| Posterior circulation ischemic stroke | 1 (0.02%; 95% CI, 0.00–0.10) | 8 (0.14%; 95% CI, 0.06–0.27) |
| All-cause Deaths | 56 (0.96%; 95% CI, 0.73–1.25) | 404 (6.93%; 95% CI, 6.29–7.61) |
| Intracerebral hemorrhage | 11 (0.19%; 95% CI, 0.09–0.34) | 17 (0.29%; 95% CI, 0.17–0.47) |
| Composite: ischemic stroke or death | 123 (2.11%; 95% CI, 1.76–2.51) | 546 (9.36%; 95% CI, 8.63 – 10.14) |

### Outcomes at 45 days-4 years

During the long-term follow-up window (day 45 through 4 years) shown in **Table 2**, the composite rate of new ischemic stroke or death was 9.36% (95% CI, 8.63–10.14; 546 events). Ischemic stroke of any type occurred in 179 patients (3.07%; 95% CI, 2.64–3.55), while all-cause death was the dominant component of the composite (404 events; 6.93%; 95% CI, 6.29–7.61). Anterior circulation strokes occurred in 34 patients (0.58%) and posterior circulation strokes in 8 (0.14%). Intracerebral hemorrhage remained rare (17 events; 0.29%). In the CREST-2 trial, the 4-year primary composite event rate was 2.8% (95% CI, 1.4–4.3) in the CAS group shown in the **Table 3**.

**Table 3.** Summarizes the key outcome comparisons between the LLM-driven real-world estimates and the published CREST-2 trial endpoints.

| Outcome | LLM-Derived EHR (N = 5,831) | CREST-2 CAS (N = 616) |
| --- | --- | --- |
| Periprocedural stroke or death (0–44 d) | 2.11% (1.76–2.51) | 1.3% (0.6–2.5) |
| New ischemic stroke (any) (45 d–4 y) | 3.07% (2.64–3.55) | 2.8% (1.4–4.3) |

### Protocol Extraction Performance

To evaluate protocol extraction performance (RQ1), we defined a gold-standard checklist of 26 sub-components across the five-target trial emulation design parameters derived from the CREST-2 protocol and assessed LLM outputs prior to human correction.

**Table 4** presents detailed extraction accuracy by design parameter. The pipeline achieved 92.3% first-pass accuracy (24/26 elements) with precision 1.00, recall 0.923, and F1-score 0.960. Four parameters were extracted with perfect accuracy, eligibility criteria (age ≥18, stenosis ≥70%, qualifying CEA/CAS procedure, recent stroke/TIA exclusion, prior revascularization exclusion, ≥1-year baseline enrollment), treatment strategies (CEA/CAS CPT definitions, treatment assignment, index date), time-zero assignment (procedure date, 0–44 day periprocedural window, 45 day– 4 year follow-up), and follow-up/censoring (follow-up windows, blanking period, censoring rule, time-to-event). Two errors occurred in outcome definitions (anterior/posterior stroke codes, ICH, ischemic stroke prefix, stroke-any definition, death, composite endpoint, inpatient restriction), both were syntactically valid but clinically incorrect. Each was corrected in a single human-in-the-loop iteration, indicating rapid refinement and highlighting outcome phenotyping as the most error-prone stage in LLM-generated pipelines. The perfect precision (1.00) indicates that the LLM did not hallucinate eligibility criteria, outcome codes, or follow-up rules beyond what the CREST-2 protocol specified.

**Table 4.**
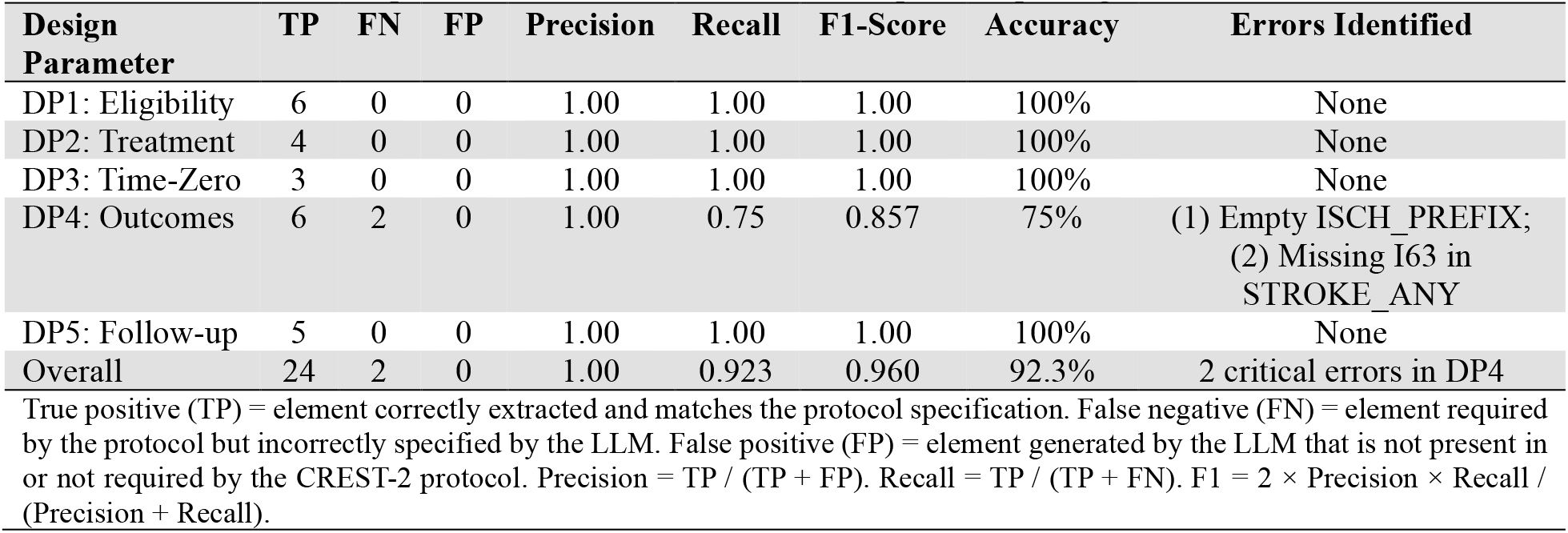
Protocol extraction performance metrics for LLM-driven protocol parsing.

### Pipeline Outcome Performance Evaluations

Further, to evaluate the performance of the LLM-driven pipeline (RQ2), we compared the patients-level outcome estimates generated from the EHR cohort (N=5,831) against the published endpoints from the CREST-2 CAS arm (N=616). Because the two populations comprise different patients from different data sources, patient-level agreement metrics are not applicable. Instead, we employed four population-level concordance metrics that are standard in comparative effectiveness research and phenotype validation studies. **Table 5** presents the complete concordance analysis across all outcome endpoints. The periprocedural composite of stroke or death within 44 days was 2.11% (95% CI, 1.76–2.51) compared with 1.3% in the CREST-2 CAS arm, with a negligible standardized mean difference (SMD = 0.058), overlapping confidence intervals, and a non-significant z-test (p = 0.138), indicating statistical compatibility. Long-term ischemic stroke rates were closely aligned with trial results (3.07% vs 2.8%; SMD = 0.016; p = 0.722), suggesting strong concordance between the LLM-derived estimates and adjudicated trial outcomes. The strongest agreement was observed for all-cause mortality (6.93% vs 7.1%; SMD = 0.007; p = 0.872), reflecting the reliability of mortality capture in EHR data and supporting the validity of the pipeline’s data extraction and temporal logic. Overall, these findings demonstrate that the automated framework produces real-world outcome estimates that are quantitatively consistent with the CREST-2 trial.

**Table 5.** Complete concordance analysis across all outcome endpoints.

| Outcome | EHR<br>(95% CI) | CREST-2<br>(95% CI) | SMD | O/E | CI<br>Overlap | z-test p | remarks |
| --- | --- | --- | --- | --- | --- | --- | --- |
| Periprocedural stroke/death (0–44 d) | 2.11%<br>(1.76–2.51) | 1.3%<br>(0.6–2.5) | 0.058 | 1.62 | Yes | 0.138 | Compatible |
| New ischemic stroke (45 d–4 y) | 3.07%<br>(2.64–3.55) | 2.8%<br>(1.4–4.3) | 0.016 | 1.10 | Yes<br>(within) | 0.722 | Compatible |
| All-cause mortality (4-year) | 6.93%<br>(6.29–7.61) | 7.1% | 0.007 | 0.98 | Yes | 0.872 | Concordant |

SMD, standardized mean difference; O/E, observed-to-expected ratio; CI, confidence interval. SMD < 0.10 indicates negligible difference; O/E 0.80–1.20 indicates good concordance with the trial benchmark; overlapping 95% CIs indicate statistical compatibility; z-test $p > 0.05$ indicates no statistically significant difference.

## Discussion

Our findings showed that LLMs can extract and operate valid TTE design parameters from published protocols. The RAG-augmented protocol parser successfully identified eligibility criteria, treatment strategies, time-zero assignment, outcome definitions, and follow-up specifications. These extractions were accomplished through structured prompting without fine-tuning, demonstrating that general-purpose LLMs possess sufficient medical knowledge and coding logic to translate clinical protocol language into executable analytic code. However, the human-in-the-loop validation phase revealed that LLM-generated clinical logic is not error-free. Two critical errors were identified during the analysis: (1) an empty ischemic stroke ICD-10 prefix that would match all diagnosis codes, and (2) an incomplete stroke-any definition that excluded ischemic strokes entirely. Importantly, both errors were syntactically valid code and would have been executed without runtime errors while producing clinically incorrect results. This highlights a fundamental challenge of LLM-generated research pipelines, errors may be logically coherent yet clinically invalid, and therefore detectable only through expert review. The proposed structured multi-checkpoint architecture, with clinician validation of clinical logic and informatician validation of technical implementation, proved essential for ensuring pipeline validity. Accordingly, the current paradigm is best described as human-supervised autonomous generation, where the LLM generates pipeline logic while domain experts ensure clinical and coding validity. A key architectural contribution of this framework is the structured five-design-parameter (5-DP) template, which serves as an intermediate representation between protocol text and executable code. The template ensures that all essential TTE components are explicitly defined before pipeline generation. Furthermore, this architecture ensures that no protected health information leaves the institutional environment, providing a practical pattern for HIPAA-compliant LLM deployment in clinical research. The study also followed a detailed ICD-10-CM classification framework for ischemic stroke based on vascular territory. This study has several important limitations that should be considered when interpreting the results. Firstly, the framework was evaluated using a single trial (CREST-2) and a single EHR data source. Secondly, LLM outputs are inherently non-deterministic, and exact code generation may vary across runs without strict version control. Lastly, the analysis did not implement randomization or confounding adjustment, and therefore comparisons with trial results are descriptive rather than causal. An important future direction is the development of automated validation mechanisms that reduce reliance on manual expert review. Potential approaches include: (1) cross-referencing LLM-generated code sets against established phenotype libraries such as PheKB, and eMERGE; (2) automated unit testing of temporal logic and code-set membership; (3) LLM-as-judge frameworks where a second model evaluates the clinical plausibility of the first model’s output; and (4) simulation-based validation where synthetic EHR data with known ground-truth event rates are used to benchmark pipeline accuracy.

## Conclusion

This study demonstrates the feasibility of an LLM-driven framework for automating target trial emulation from clinical protocols. Using the CREST-2 trial as a case study, the system extracted core trial design parameters, generated executable phenotyping pipelines, and produced real-world outcome estimates from EHR data that were quantitatively concordant with trial-reported results. The framework introduces three key contributions: a structured five-design-parameter template, a multi-model architecture that enables HIPAA-compliant processing by separating protocol reasoning from patient-level computation, and a vascular territory–specific stroke classification scheme that improves outcome granularity in claims-based analyses. Human-in-the-loop validation also confirmed that expert oversight remains essential for ensuring clinical correctness. Future work will evaluate the framework across multiple trials and data models, integrate automated validation mechanisms, and incorporate causal adjustment methods. Together, these advances provide a foundation for scalable, AI-augmented real-world evidence generation that complements clinical informatics expertise.

## Data Availability

The data used in this study were obtained from the Oracle Health Real-World Data (OHRWD) repository and are subject to strict data use agreements and privacy regulations. The authors are not authorized to share these data. Researchers may obtain access through Oracle Health subject to appropriate approvals. The analytical code and methodology are available from the authors upon reasonable request.

